# Perspectives and experiences of consumers and bicultural health educators on the diagnosis and management of asthma

**DOI:** 10.1101/2025.06.15.25329655

**Authors:** Mahisha Jayakody, Esmerelda Rivas, Khan Vinh Nguyen, Sarita Singh, Hoan Tran, Bodil Rasmussen, Lata Jayaram

**Author notes:** Corresponding author: Lata Jayaram Department of Respiratory and Sleep Medicine Western Health, 160 Gordon St, Footscray, Victoria 3011.

## Abstract

**Purpose:** Lack of access to culture-specific asthma care contributes to poor asthma outcomes in Culturally and Linguistically Diverse communities. A nurse-led asthma education programme incorporating Bicultural Health Educators to improve asthma outcomes was trialled in three of the communities most severely affected during a thunderstorm asthma event in West Melbourne, Victoria, Australia. This study aimed to uncover unique asthma management challenges affecting these communities.

**Patients and Methods:** A total of 47 participants with asthma from Vietnamese, Sri Lankan, and Indian communities completed two asthma education sessions, six weeks apart, with an Asthma Nurse Educator supported by a Bicultural Health Educator. At the first session the participants participated in a questionnaire and a narrative interview in which they shared their views on their asthma healthcare journey. Bicultural Health Educators recorded responses to open questions about perspectives and experiences of their journey, which were coded and thematically analysed.

**Results:** Key factors impacting on asthma management from the consumer and Bicultural Health Educator perspective included limited knowledge of asthma, limited understanding of asthma management, difficulties with inhaler technique and difficulty navigating asthma care in the community. Culturally-driven attitudes towards asthma management, along with low educational literacy levels in some consumers also played a significant role.

**Conclusion:** The study identified factors impacting on asthma management in the Sri Lankan, Indian and Vietnamese communities in West Melbourne. Incorporating these insights into the delivery asthma care and education might improve asthma outcomes in these populations.

## Introduction

People of culturally and linguistically diverse (CALD) communities face barriers to accessing healthcare at the patient, clinician and health system level. Australian and Internation data indicate that people from racial and ethnic minority populations have lower primary care attendances, but significantly higher rates of secondary healthcare utilisation due to asthma, [1] and are frequently reported to have poorer asthma outcomes. [1,2] This was reflected in local hospital data from Western Health, with CALD people with asthma having lower levels of attendance to outpatient follow up and lung function testing appointments compared to the general population, which may be contributing to poor asthma outcomes in CALD consumers. Furthermore, during the epidemic Victorian thunderstorm asthma event in 2016, patients of Indian, Sri Lankan and South-east Asian origin represented 39% of the asthma-related emergency department presentations, and accounted for six of the ten deaths that occurred; 79% of these patients were born overseas. [3]

The provision of cultural support via multicultural health workers has been studied in both Australian and international settings as a strategy to help CALD patients navigate barriers in accessing healthcare. [4,5] Compared to usual care, culture-specific asthma education programs have been identified to result in a greater improvement of asthma quality of life in both adult and paediatric populations and greater reductions in the rates of severe asthma exacerbations in children.[2] We are trialling a nurse-led asthma education program, in which culture-specific asthma education sessions are provided to participants from Vietnamese, Indian and Sri Lankan communities in the Western suburbs of Melbourne with the support of Bicultural Health Educators (BHEs) to improve their asthma outcomes. The aim of this study was to uncover perspectives from consumers of the three communities about asthma and its management.

## Methods

A qualitative explorative design [6] with individual narrative interviews was applied to gather the perspectives and experiences of participants and BHEs.

### Participants

A total of 47 adults greater or equal to 18 years of age living with asthma were recruited, using convenient sampling from 3 communities: Sri-Lankan (Sinhalese) (n=8), Indian (Hindi) (n=24) and Vietnamese (n=15) between March 2023 to March 2024. Participants were recruited from respiratory outpatient clinics, following asthma-related hospital admissions, GP clinics and directly from the community through advertising and reaching out.

### Procedure

Participants received an initial asthma education session led by an asthma nurse educator and a Bicultural Health Educator. This included a baseline questionnaire that collected their demographic data and assessed their initial understanding of asthma and its management. They were then invited to participate in an exploratory narrative interview about perspectives and experiences in managing their asthma. This was followed by 1-2 hours of asthma education program that discussed asthma pathophysiology, symptoms, triggers, treatment, inhaler technique and self-management skills using the teach-back method. The education was reinforced in a 6-week follow-up session either in person on site, or via telehealth. Both sessions were led by an asthma nurse educator and supported by a Bicultural Health Educator (defined on page 3). The Bicultural Health Educators documented participants’ thoughts, ideas and perspectives following interviews.

### Data analysis

Descriptive analysis was used to describe baseline demographics and quantitative data was presented as frequencies. BHE documented interviews in notes during and after the interviews. The documentation from each interview was analysed using content (conceptual) analysis to identify the presence and meaning of certain themes or concepts.[7] The interview documentation was de-identified and was repeatedly read by members of the research team (MJ, LJ, SS,CV,) and was grouped into meaningful themes to explain and interpret the content from the interview documentation. The identified themes were discussed among the research team (all authors) until a consensus was reached.

## Results

Participant baseline characteristics are outlined in Table 1. Approximately two-thirds of the participants were female. Over 46% were aged between 40 to 59 years and over 59% had a diagnosis of asthma for at least 5 years. 14% of participants were illiterate in their own language or had not completed school. Most had a nominated GP but did not see them regularly.

**Table 1.** Baseline characteristics of participants.

|  |  |
| --- | --- |
| **N** | **47** |
| **Gender** |  |
| Male | 15 |
| Female | 32 |
| Other | 0 |
| **Age range, years** |  |
| 20-39 | 9 |
| 40-59 | 22 |
| 60+ | 16 |
| **Ethnicity** | 15 |
| Vietnamese | 8 |
| Sri Lankan | 24 |
| Indian |  |
| **Duration of Asthma** |  |
| < 5 years | 19 |
| 5 -15 years | 24 |
| > 15 years | 4 |
| **Highest level of education** |  |
| did not attend school; low educational literacy grade 1a or less (can't read or write in own language) | 7 |
| completed high school | 16 |
| TAFE/Polytechnical school | 5 |
| University | 9 |
| did not want to answer | 10 |

### 1. Limited knowledge of asthma

Various levels of asthma knowledge among participants were BHEs identified. Limited understanding of basic asthma pathophysiology and symptoms was a common finding among participants from all educational levels. Definitions of asthma were often erroneous and sometimes based on spiritual concepts (Table 2).

**Table 2.** An overview of participants’ responses to the question “What is asthma?” during the baseline questionnaire.

|  |  |
| --- | --- |
| **N** | **42** |
| Difficulty breathing | 6 |
| Airway constriction | 3 |
| Cough | 3 |
| Swollen lungs | 1 |
| Allergies-related condition | 9 |
| Weather related condition | 1 |
| Suffocation | 4 |
| “Wind” | 2 |
| A foreign body obstructing airway | 1 |
| Oesophageal constriction | 3 |
| Spiritual concept | 4 |
| Don't know | 9 |
| Did not wish to answer | 1 |

When asked “what is asthma” (defined as episodic wheeze, chest tightness or breathlessness for this study), 80% felt they did not have a good understanding of what asthma was; 19% answered “don’t know”, one attributed it to a foreign body in the airway, and three attributed it to oesophageal constriction. When asked about understanding of asthma, Participant 12 responded:

> *“I don’t know, I was told that I got asthma, but I don’t have any understanding of the disease. The only thing that I know is that I struggle to breathe and to do any form of activity. For example today, I walked from the car to this office and I feel out of breath.”*

Many participants were not aware of their asthma triggers; participant 8 was *“not aware that winter or changes in season can be a trigger for Asthma”* and participant 25 *“was uncertain…whether he was allergic to pollen”*

Some participants did not recognise asthma to be a chronic disease requiring symptom control through lifelong therapy and had expectations of being cured. Participant 4 expected it will *“resolve within a couple of years”,* Participant 19 was “*really confused why he is getting asthma again, thought he was cured when he was given some traditional medicine when he was a child back in Vietnam.”* Consequently, an improvement in symptoms or a reduction in medication use was not recognised as a sign of progress; any presence of symptoms or inhaler requirement was viewed as being unwell.

### 2. Limited understanding of asthma management

#### 2.1 Difficulties in using asthma medication appropriately

Perspectives and experiences from both participants and BHEs demonstrated that the majority did not know the difference between a preventer and reliever, could not identify the correct time to use each medication, and could not distinguish the different inhalers from each other (Figure 2).

**Figure 1:**
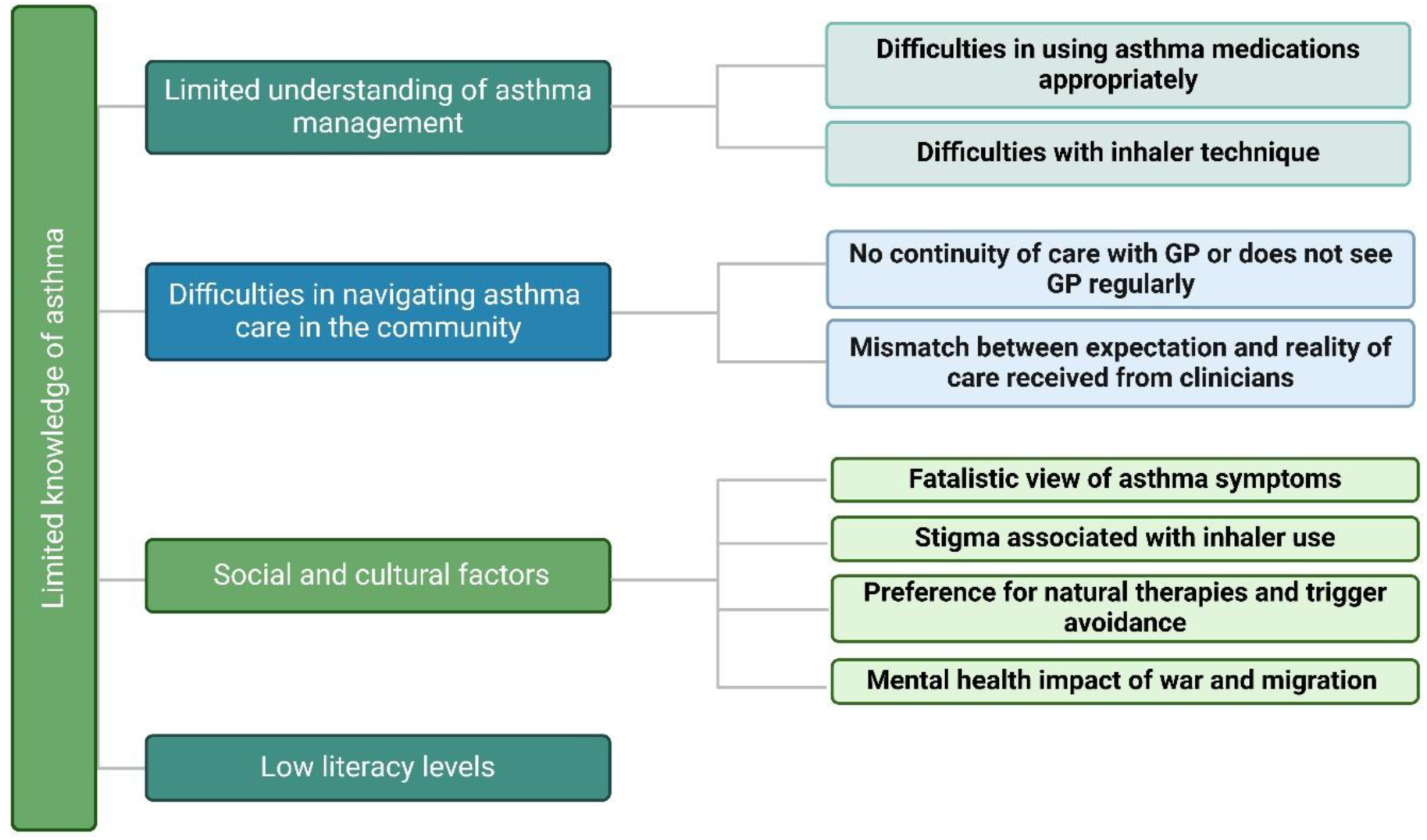
Factors impacting asthma management among participants, as identified by participants and BHEs.

**Figure 2:**
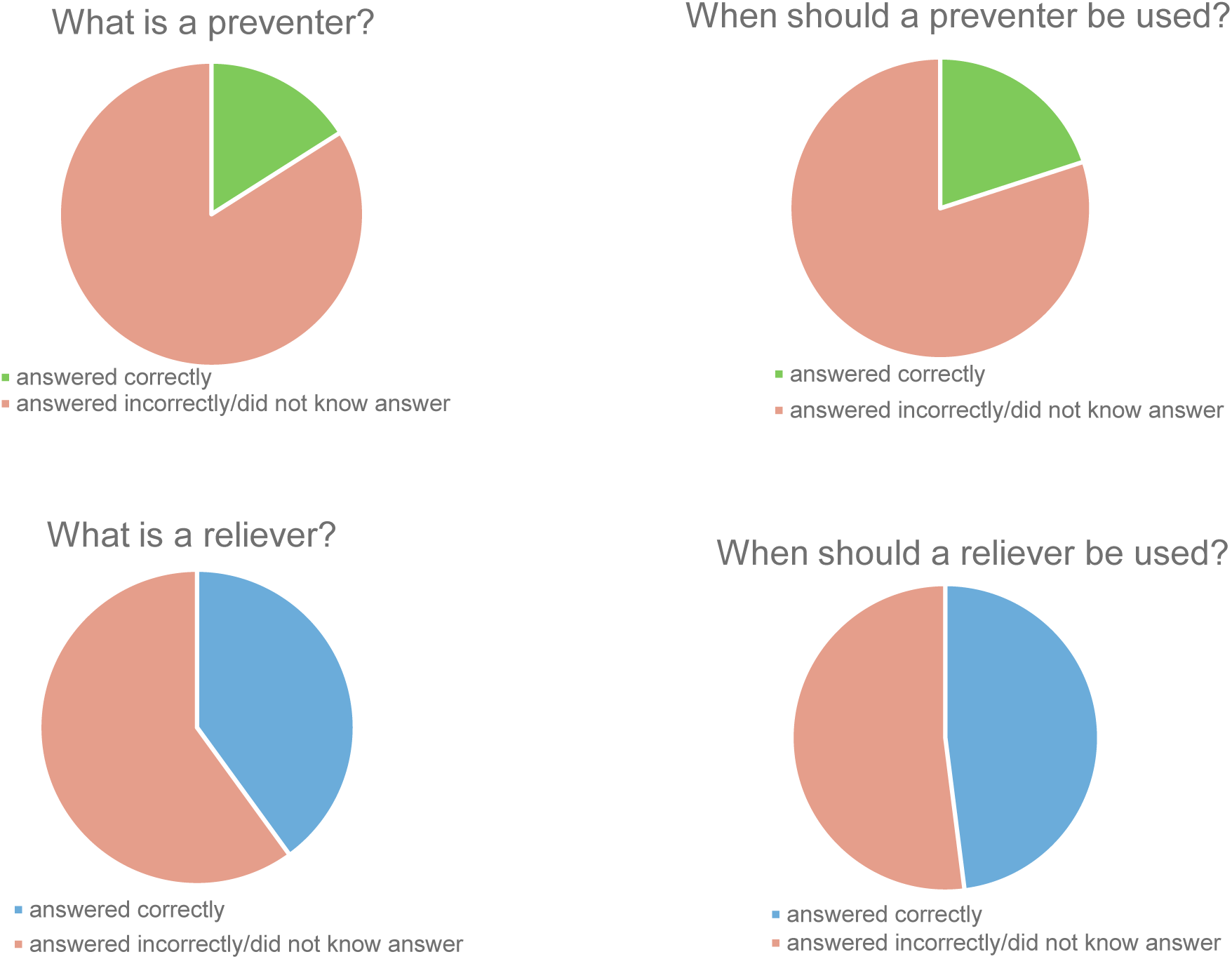
Participant knowledge of asthma relievers and preventers as per baseline questionnaire

**Figure 3:**
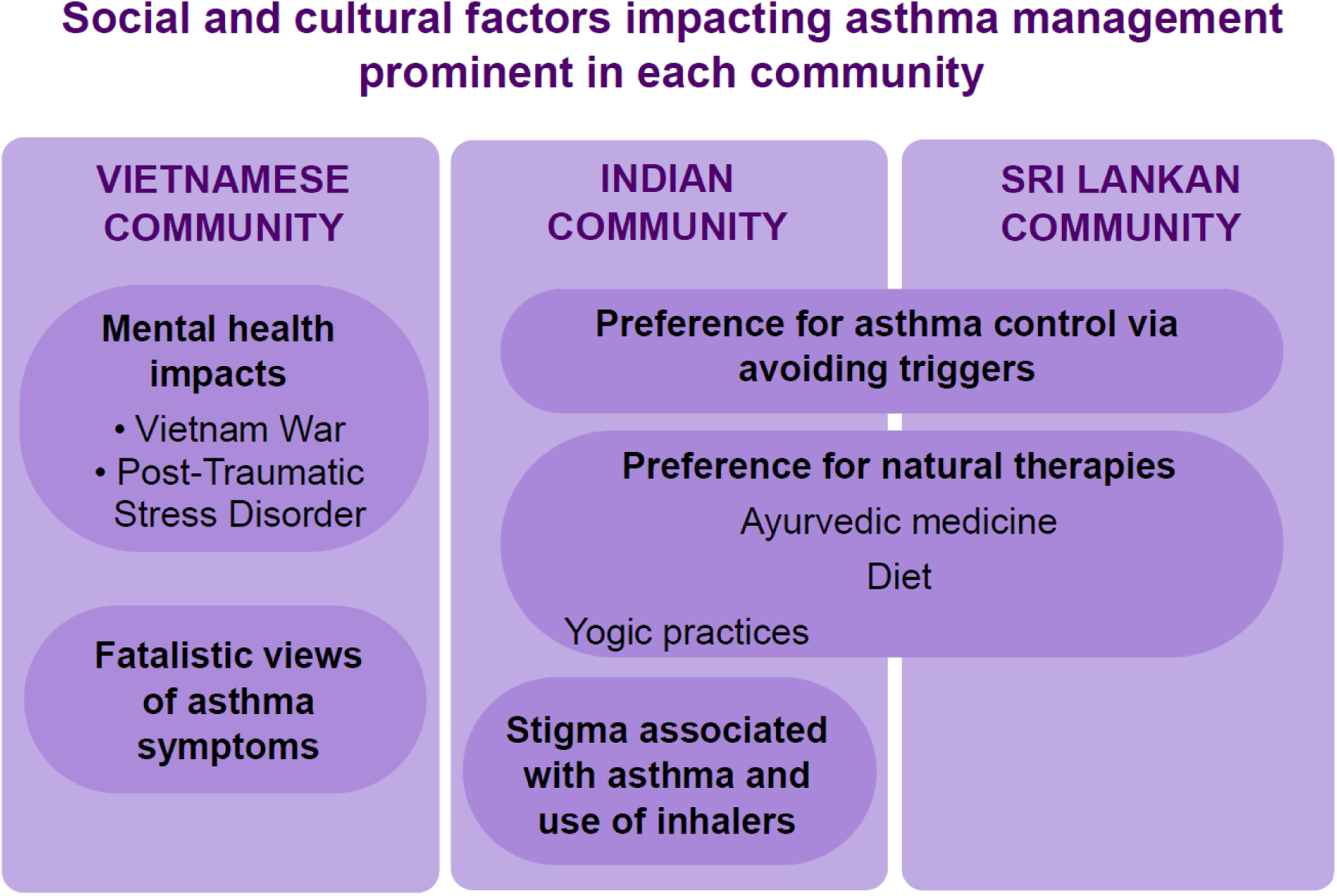
Social and cultural factors impacting asthma management prominent in each community.

Some participants did not understand the need for preventer use when they were well, and only took preventers when symptomatic. Participant 9 *“said that no-one has never mentioned or described the actions of her Symbicort inhaler…she did not see the need to use her regular medication with no obvious symptoms.”* Most participants used their reliever regularly, used it instead of a preventer, or overused it.

Many participants did not know how to escalate their asthma therapy during exacerbations; for instance, Participant 12 *“brought medication with him - they include Atrovent [ipratropium], Ventolin [Salbutamol], Pulmicort turbuhaler [Budesonide], Spiriva [Tiotropium bromide], Symbicort [Budesonide/Formoterol] and prednisolone. When he is having an asthma attack, he will take one or two puffs from each of the above medication.”*

Many participants also did not know how to track their medication use or read the built-in dose counter on the inhaler, and as a result continued to use empty inhalers. Participant 11 “*didn’t know when to tell if device was running out. He has never paid attention to the number/colours on puffer.”* Participant 16’s *“Ventolin expired in 2020”* as he was “*not aware where to find expired date on puffer”.* Multiple participants had not visited a GP to get up-to-date prescriptions to replace their medications, including Participant 18 who *“has been using her friend’s reliever puffer*..*.”*

Notably, no participants reported possessing a current written asthma action plan which they understood. Some participants did not recognise a need for dedicated asthma education as they perceived themselves to have adequate knowledge of asthma through lived experience; Participant 2 *“declined an asthma plan, stating that “I have been asthmatic all my life, I think I know what to do.”* On further exploration, there were significant knowledge gaps identified in these participants, which were addressed by the BHEs. One BHE observed that Participant 11, who had errors in spacer and inhaler technique *“never rinsed or cleaned his spacer…was not aware of how to use his spacer without holding his breath. He felt that he knew everything as he attended GP and hospital clinic appointments regularly*.”

### 2.2 Difficulties with inhaler technique

Some participants had no previous opportunities to develop their inhaler technique, as no one had spent time explaining inhaler technique to them. Participant 14 *“did not realise there is a correct and incorrect technique.”* BHE 2 observed: *“Because they have been using medication for a while they thought they were confident in what they were doing and accepted that their symptoms were [their] baseline, [and] not because they were not getting the medication administered correctly.”*

Common issues noted in inhaler technique were improper mouth seal with the spacer or inhaler mouthpieces and not taking in the adequate number of breaths between puffs. As observed by the BHEs, Participant 5 demonstrated *“4 to 6 puffs one after the other without holding breath in or a break between puffs”* and Participant 13 *“placed her puffer at the tip of her mouth, and delivered one puff into the air and breathed in and out 3 times into the puffer.”*

Some participants were unaware of the need for a spacer, and some lacked knowledge around spacer technique and maintenance. Participant 19 *“was not aware that you are allowed to breathe your waste air (or exhale) back into the spacer.* Participant 13 *“demonstrated her technique using the spacer - she delivered 5 puffs continuously and then took one big breath in.”* Similar to participant 11, participant 7 “*didn’t have knowledge about cleaning spacer regularly or when dirty”*

### 3. Difficulties in navigating asthma care in the community

While most participants had a nominated GP (Table 1), they did not seek regular care or did not have continuity of care with a single GP. BHE 1 and 2 observed:

> *“A lot of patients do not understand the importance of longevity of care by the GP. This should be explained to patients and highly encouraged.”*

BHE 1 observed:

> *“Patients are seeking care from health centres but they 1) do not rebook patients with same GP and 2) do not allow patients to ask more than one question/problem in the consult. This is a huge problem because they are not dealing with patients as a whole and can often miss diagnosing or attending to things that need to be looked into.”*

Participants reported instances of attending the GP clinic when feeling very unwell with asthma, which resulted in them being sent to ED, which led to the perception that GPs cannot provide asthma care when they are unwell. However, as noted in previous sections, limited knowledge about escalating asthma therapy during an exacerbation may result in presentations to the GP at a more unwell stage that cannot be adequately managed in the GP setting and warrants emergency care. For example, participant 3 “*expressed feelings of frustration with his GP. When he attended GP, he felt very unwell, unable to breathe and he was told by GP that he could not give him oxygen as it’s too expensive, ambulance was called (participant story telling).”* This experience negatively impacted his trust in his GP, making him less likely to seek early GP care for subsequent exacerbations, and more likely to present directly to ED.

Participants noted challenges to obtaining asthma education during clinic or GP visits. Participant 3 expressed *“his discontent as there is no education given about his condition.”* Participant 23, who was taking inhalers and tablets for asthma, expressed a need for *“more information on side effects of long-term medications”* Additionally, participants felt that clinicians did not accommodate for the challenge of understanding medical terminology coupled with a language barrier; participant 17 reported that while her doctor provided information on asthma, she *“did not comprehend this and would like someone to explain to her in plain English.”* BHE1 observed:

> *“Patients are eager to speak in English. But they cannot understand everything in English and their confidence may not be a true representation of their comprehension. [They] should have a support worker, especially if they (workers) have health literacy because this can help with comprehension. i.e. health background.”*

### 4. Asthma in the context of cultural and social experiences and beliefs

When discussing poor preventer usage, participant 9 reported that “*in her culture, medication is only used when someone is really sick, if there is no symptoms medication is disregarded.”*

Experiencing shortness of breath from asthma carried various social and spiritual connotations in some cultures. Participants in the Vietnamese community held fatalistic spiritual views regarding the symptom of breathlessness; Participant 12 expressed the belief that “*your breath is your life and once it is gone you are dead, so this increases the fear of dying from asthma.”* Each asthma exacerbation was perceived as a loss of ‘vitality’ or ‘life essence’ and a sign of inevitable death which could not be influenced by medical treatment.

Some Indian participants expressed that there was social stigma associated with using inhalers, with 42% therefore feeling reluctant to use inhalers regularly. As one BHE noted:

> *“In India, people often call asthma “Dama”…Participant 3 said her mother told her, “Do not tell anyone in the family, relatives, or friends that you have Dama.” This advice comes from worries that sharing this information could hurt her chances of getting married. People might see her as unhealthy or imperfect and question her ability to have children. Because of this, she stopped using her inhaler in public and looked for places to hide when she needed to take puffs.”*

Participants from Indian and Sri Lankan backgrounds showed an interest in controlling their asthma through methods they believed to be more “natural” with less side effects, including dietary modifications, avoidance of triggers, yoga and traditional or *Ayurvedic* therapies, often preferring these to inhaler therapy. Participant 9 was advised by her friend *”Steroids are addictive and have more side effects, which cause more harm than good.”*

BHE 2 noted:

> *“Indian participants often select homeopathy or Ayurveda for chronic conditions, viewing these methods as safer over time…Patients often receive Ayurvedic, homeopathic, or herbal supplements from their home country, sometimes without their allopathic doctors knowing.*

*This strong belief in Ayurvedic principles is a key part of healthy living for Asian Indians, taught to them from a young age.”*

Participant 23 had *“previously seen an Ayurvedic practitioner in Sri Lanka for her chronic cough, and did not think her cough was connected to asthma”*.

The hot and cold theory of illness was a prominent belief within the Indian and Sri Lankan participant groups, in which certain foods were attributed the property of being “hot” or “cold”; consuming “cold” foods were believed to exacerbate mucus production and airway disease, and consuming “hot” food was believed to result in inflammation. Participant 13 believed that *“eating cold food makes it hard to breathe and causes coughing; otherwise, I have no disease*”. As one BHE observed, *“They think allergies are due to an imbalance of hot and cold and that it’s important to avoid all foods that are considered “cold,” like rice, yogurt, ice cream, salads, juice, etc.”*

Many older participants placed the onus on health professionals to provide all necessary information on asthma management, due to attitudes built through interacting with more paternalistic healthcare systems in their countries of origin. They were inclined to accept their diagnoses without seeking further information. Participant 12 *“[did] not have knowledge of asthma; it is something that they have just been told and accepted.”* Participant 13 was *“Dependent on healthcare professional - ‘do as told’.”*

For many of the participants, the acculturative stress of migration had created a profound sense of displacement, which affected their asthma management experience and their interactions with the healthcare system. This was especially true for the Vietnamese population, which included those who had migrated to Australia as refugees during the Vietnam war. Participant 13 *“described having asthma since her younger years when placed on a concentration camp, but as the years progress [felt that] her asthma is not well controlled.”* 45% of the participants reported stress, anxiety or depression as having an impact on their asthma management – of these, 53% were from the Vietnamese community. BHEs observed that the mental health needs of some participants who suffered from post-traumatic stress following the Vietnam war had not been adequately addressed, and over time led to issues with substance misuse and breakdown of family and community relationships. This resulted in social isolation and feelings of intense anxiety around being unwell from asthma; for example, participant 12 “*elaborated on [his] fear of dying from an asthma attack and that he does not have close relatives to check on him.”* For this participant, the burden of mental illness was a barrier to regular medication adherence and attending appointments. Memories of conflict experienced in his home country was also a trigger for shortness of breath, which he attributed to asthma and would attempt to treat with inhalers. He had never received formal mental health services, and was not equipped to identify or manage physical symptoms of anxiety including panic attacks.

Family and personal stressors were also noted to influence health behaviours. Participant 6 described *“personal/family issues and how this is a trigger for her asthma and her smoking…She was very honest about her smoking habit and how hard it is for her to quit.”* Participant 15 reported that her struggles with mental health made it difficult to adhere to consistent asthma medications use, but felt *“reluctant to deal with mental health because of social stigma, cost*.”

### 5. Impact of language literacy on asthma management

Low literacy levels in both English and first languages was noted in fifteen per cent of participants, as evidenced by self-report and targeted questions on the baseline questionnaire, (Table 1) making it particularly challenging for them to obtain asthma information. Participant 13 was from a *“Low education background from a rural area in Vietnam”* and “*stated that she can’t read or write and this makes things a bit harder for her. She was apologetic about it, as she believes this interferes with her level of comprehension to follow instructions.”*

Low literacy levels also affected the participants’ ability to advocate for themselves and to seek further understanding of their diagnosis, as noted by BHE1:

*“Their low literacy/academic experience lessens their confidence to feel empowered in their diagnosis. They start the session with little confidence about understanding asthma and accepts this without trying to seek understanding. Once they were educated in a way that is understandable to them, it is clear that there is a shift in their confidence and becomes more positive regarding their outcome. Fear of dying from asthma is settled.”*

Participants often responded well to analogies and explanations related to their BHE1 observed that *“It is very helpful to ask the patient what they do/did for work. This can help with explanations for the lay person e.g. a handy man can have airways described as pipes.”*

## Discussion

This study describes the complex challenges faced by culturally and linguistically diverse communities in managing their asthma, focusing on three cultural groups (Vietnamese, Indian and Sri Lankan) who were most severely impacted by the 2016 Thunderstorm Asthma event in Melbourne. [3,8,9] While this event has been widely studied in the literature. This paper, to our knowledge, is the first that explores specific, persistent challenges to long-term asthma control in these affected groups from a consumer perspective.

The limited knowledge of asthma among the participants was a significant barrier to adequate asthma management, and may be due to the fact that asthma is a relatively unfamiliar disease within the migrant populations featuring in this study. Interestingly, multiple Australian studies have demonstrated that South Asian and East Asian migrants who have no allergic or atopic symptoms pre-migration, showed an increased susceptibility to asthma post-arrival in Australia compared to the general Australian population. [9–12] This is thought to be due to sensitisation to common environmental triggers such as grass pollen over time, with the incidence of asthma directly correlating with individuals’ length of stay in Australia.[8,10,11] It is unsurprising that many participants in this study had limited or no knowledge of asthma at the time of their diagnosis. The unfamiliarity with asthma is perpetuated by language barriers that prevent self-education using the available (predominantly English) asthma resources, making this vulnerable group ill-equipped to handle their diagnosis without significant support from their clinicians.

Some participants, particularly those of a low educational background, showed a tendency to accept information provided by their healthcare professionals without seeking additional clarification. This attitude appears to be a complex combination of lack of empowerment within an unfamiliar culture and health system,[13–15] as well as being raised in cultures with a more paternalistic healthcare system where patients would often yield to physicians in making treatment decisions without ‘questioning the authority’ of the physician.[16] Identifying and addressing these factors is an important part of empowering patients to express their concerns and expectations, as demonstrated by the BHEs in this study.

With regards to asthma management, our data suggested that while the role of relievers was widely understood, the role of preventers was less known. This was often accompanied by the belief that asthma is an acute condition rather than chronic, and the condition is only present when symptomatic. This is similar to the findings of a survey conducted in Singapore, [17] where 62% of participants felt that the use of relievers was the best way to manage their asthma; 28% relied solely on a reliever and preferred it over their preventer. A study with South Asian participants with asthma in the UK observed a lack of familiarity with preventer inhalers and asthma action plans amongst first-generation and recent migrants, also noting that reactive and curative care, rather than preventative care, is the norm in their countries of origin. [18] Social stigma around inhaler use and a reluctance to use medication unless visibly symptomatic were significant factors in poor preventer adherence. [13] The preference for alternative methods to control asthma among South Asian patients, such as avoidance of certain foods and practising yoga, has been noted in studies [13, 18] and was reflected among the Indian and Sri Lankan participants in our cohort.

For many participants of this study, poorly controlled asthma had a restrictive impact on mental health, social life and physical activity. This is congruent with findings in previous literature; for example, 55% of adults surveyed in Singapore found asthma to have limited their physical activity in the last month, and 42% felt low in mood because of their asthma. [17] Mental health issues were noted to influence asthma control for some participants in this study, and in the cases of some Vietnamese participants, were compounded by post-traumatic stress and social isolation. Managing the complex mental health overlay to asthma care in these cases requires cultural support as well as referrals to appropriate mental health services, along with community de-stigmatisation of the diagnosis and seeking treatment. The presence of BHEs is particularly valuable in uncovering such issues, which consumers are unlikely to disclose to their clinicians during routine care.

The use of “language-concordant care”, which refers to the physician being fluent in a patient’s preferred language [19], has been demonstrated to be helpful when delivering complex self-management education to CALD patients in hospital and community settings; One study showed significant improvements in glycaemic control among limited-English-proficiency Latino patients with diabetes who switched from language-discordant to concordant Primary Care Physicians. [20] Another study demonstrated that providing discharge education through a language-concordant physician is associated with reduced 30-day readmissions. [21] Within our cohort, the use of language-concordant GPs was a prevalent strategy. Studies have also demonstrated the benefit of multicultural health workers in improving health behaviours in culturally and linguistically diverse communities. [5,22] Here, Bicultural Health Educators operated as a form of community-based language concordant care. The self-management skills required for optimum control of asthma may be initially complex to teach; as such, the high level of health literacy among our BHEs ensured that they were equipped to provide tailored asthma education according to the education, occupational and cultural background of the consumers, which in turn aims to empower them in managing their disease.

Hearing directly from of consumers navigating the health system with asthma is a key strength of this study. Only three communities’ perspectives, however, are documented and therefore these results may not be generalisable. Given that some participants were recruited from respiratory clinics and following hospital admissions, the study may be capturing a cohort with a higher prevalence of severe asthma compared to each community. While some of the perspectives identified in this study may apply to other CALD communities, they may have their own barriers to care, which needs to be explored though further research.

### Implications for future practice

The reflective nature of this of this study provide insight into unique cultural experiences and perspectives on asthma management seen among the three (Vietnamese, Indian and Sri-Lankan) communities, and provide a roadmap for structuring future Bicultural Health Educator-supported asthma education sessions to better fulfil the needs of these consumers.

## Conclusion

This study highlights the complex barriers to asthma management in the Sri Lankan, Indian and Vietnamese communities in West Melbourne, and emphasises the need for asthma care that considers linguistic, cultural, and individual factors. Moving forward, integrating cultural support into the provision of asthma education and asthma management may contribute to improved asthma outcomes in the above groups.

## Author contributions

All authors contributed to the study conception and design, material preparation, data collection and analysis. The first draft of the manuscript was written by Mahisha Jayakody and all authors commented on previous versions of the manuscript. All authors read and approved the final manuscript.

## Abbreviations

BHE: Bicultural Health Educator. A BHE for the purpose of this study is defined as an individual from the studied communities, who has high levels of health literacy and can represent their community and culture through lived experience.
CALD: Culturally and Linguistically Diverse
GP: General Practitioner
Participant: patient or consumer

## Data Availability

All data produced in the present study are available upon reasonable request to the authors

